# Phecoder: semantic retrieval for auditing and expanding ICD-based phenotypes in EHR biobanks

**DOI:** 10.64898/2026.01.08.26343725

**Authors:** Jamie J. R. Bennett, Simone Tomasi, Sonali Gupta, VA Million Veteran Program, Georgios Voloudakis, Panos Roussos, David Burstein

## Abstract

**Background:** Electronic health record (EHR)–based phenotyping underpins genome-wide association studies, yet current ICD-code phenotypes rely heavily on manually curated lists such as Phecodes. These definitions are labour-intensive to maintain, inherently subjective, and may omit clinically relevant diagnostic codes, reducing study power. Advances in text embedding models offer an opportunity to automate and standardize ICD-based phenotype construction.

**Methods:** We developed Phecoder, an ensemble of pre-trained text embedding models that rank ICD codes by similarity from free-text phenotype descriptions. Nine embedding models and multiple unsupervised ensemble rank-fusion methods were evaluated against 1,125 PhecodeX phenotypes. Retrieval performance was assessed using recall and average precision at top-100 (R@100, AP@100). Expert clinical review of six neuropsychiatric phenotypes was undertaken to identify relevant ICD codes absent from PhecodeX. Cohort sizes under these new definitions were compared with PhecodeX across sex and ancestry strata in the Million Veteran Program (MVP).

**Findings:** Among individual models, Qwen3-Embedding-4B achieved the highest median recall (R@100 = 0.86). Ensemble rank-fusion further improved R@100 by 3%, and median AP@100 by 8%. Expert review confirmed that Phecoder retrieved additional clinically relevant ICD codes beyond PhecodeX across all six neuropsychiatric case studies. Median potential case expansion increased by 200%, with 700% increases for bipolar disorder and 2000% increase for eating disorders.

**Interpretation:** Manually defining ICD phenotypes has been critiqued as subjective, potentially yielding overly restrictive definitions that miss relevant codes. To address this issue, Phecoder algorithmically identifies relevant codes for ICD-based phenotyping. Phecoder extracts relevant ICD codes to expand the potential case pool across different demographic groups. Phecoder is easily applicable to future ICD-code releases and across different ICD coding versions that are used in different countries. Taken together, Phecoder has the potential to improve reproducibility in EHR data research.

**Funding:** This research was supported by the Department of Veterans Affairs MVP (MVP-000, MVP-076 and MVP-096). The MVP is supported by the Office of Research and Development, Department of Veterans Affairs. The authors thank the MVP staff, researchers, and volunteers, who have contributed to MVP, and especially who previously served their country in the military and now generously agreed to enroll in the study (see mvp.va.gov for more information). The contents do not represent the views of the U.S. Department of Veterans Affairs or the United States Government.

This study was supported by the Veterans Affairs Merit grants: BX006500 (to D.B.) and BX004189 (to P.R.). This work was supported by the National Institutes of Health (NIH): R01MH125246 (to P.R.), R01AG078657 (to G.V.), R01AG067025 (to P.R.), and U24AG087563 (to P.R.).

**Research in context:** *Evidence before this study:* Phecodes are curated groupings of ICD-9 and ICD-10 diagnostic codes designed to create clinically meaningful phenotypes for large-scale research using electronic health records. They provide a standardized, reproducible way to map tens of thousands of ICD codes into interpretable disease concepts, and they are widely used in genome-wide association studies, phenome-wide association studies, and disease prediction models. PhecodeX, released in 2023, is the most recent update to this framework. It restructures the original catalogue to make full use of ICD-10 granularity and introduces more than 1,700 new phenotypes across a broad range of clinical domains. Phecodes have become foundational tools for EHR-linked biobanks, enabling harmonized phenotyping across institutions and cohorts. A major barrier to the continued development and updating of Phecodes is their reliance on slow, manual curation, which is inherently subjective. As a result, it is difficult to ensure complete capture of all clinically relevant ICD codes; particularly for conditions with diffuse presentations, heterogeneous coding practices, or evolving diagnostic criteria. Thus, some studies using predefined ICD-based phenotypes have reported unexpectedly low sensitivity, reinforcing the concern that curated code lists may miss substantial numbers of true cases.

*Added value of this study:* This study introduces Phecoder, a semantic retrieval framework that streamlines ICD-based phenotyping by ranking codes according to their similarity to any free-text phenotype description. This approach replaces static, manually curated lists with a flexible workflow in which phenotype definitions can be rapidly generated, audited, and refined simply by adjusting the input text. Phecoder therefore supports the continuous development of more comprehensive and responsive phenotype mappings. We provide the first systematic benchmark of text embedding models for ICD-level phenotyping, evaluating nine encoders and several unsupervised ensemble methods against 1,125 PhecodeX phenotypes. Leveraging existing curated mappings as a reference standard, we show that a score-level ensemble improves retrieval performance over individual models and achieves perfect median recall for mental health phenotypes. Phecoder also identifies clinically relevant ICD codes beyond PhecodeX. Expert review of six neuropsychiatric case studies confirms the clinical relevance of these additional codes, and their incorporation exposes sizable untapped patient cohorts in the Million Veteran Program.

*Implications of all the available evidence:* Together with existing use of Phecodes in major biobanks, these findings show that curated phenotype systems remain indispensable but benefit from scalable, transparent tools that support their maintenance and evolution. Phecoder enables continuous auditing and expansion of ICD-based phenotype definitions, and improves cohort completeness across demographic groups. Consequently, we anticipate that Phecoder will facilitate improved reproducibility across demographic groups in scientific research.

## Introduction

Electronic health record (EHR)-based phenotyping is foundational to modern biomedical research and healthcare delivery, supporting applications ranging from large-scale genetic epidemiology to clinical informatics, quality measurement and health systems operations ^1–4^,. These phenotypes are typically composed of diagnostic codes, namely International Classification of Diseases (ICD) codes^5^; ubiquitous across biobanks and healthcare systems.

Substantial effort has been directed towards the creation of Phecodes^6–8^: curated sets of ICD code shortlists that define thousands of clinically meaningful phenotypes. The value of Phecodes in large-scale biomedical research cannot be overstated, yet they are not without limitations. As the authors of PheCodeX have acknowledged, the manual curation of phecodes is “inherently subjective and non-quantitative”^8^. Furthermore, constructing comprehensive, reproducible phenotype definitions from tens of thousands of ICD codes is labour-intensive and vulnerable to incomplete ICD code capture, resulting in under-ascertainment in patient cohorts. For example, a recent study that compared three different manually curated ICD code lists to identify suicidal attempt phenotypes within a single cohort found substantial heterogeneity in sensitivity (30.7%-56.3%) when evaluated against survey-based C-SSRS assessments^9^. This motivates the need for systematic, scalable tools to support and accelerate manual curation of ICD-based phenotypes.

A growing body of work has explored the use of text embedding models, including those derived from large language models (LLMs) to automate aspects of EHR phenotyping. These approaches often rely on pre-trained models, which offer a low barrier to adoption by avoiding the need for task-specific model training, large proprietary corpora, or specialised computational infrastructure. Recent evaluations have shown that general-purpose, pre-trained models perform well on certain clinical phenotyping tasks^10^, and even rival EHR-specific models^11^, although others have concluded they are poor medical coders^12^; thus, it is important to evaluate their performance for specific use cases before deployment. Applications of text embedding models to phenotyping studies have primarily focused on phenotyping at the patient level^13–16^, for example, extracting phenotypes from clinical notes^15^. Beyond patient-level phenotyping, text embedding models have been incorporated into tools that support generative querying in EHR-linked biobanks, including systems that draft executable queries and phenotyping logic^17–19^. While previous work has investigated deriving ICD code embeddings from text descriptions^20^, we are not aware of any prior studies leveraging text embedding models to facilitate the automated auditing and expansion of ICD-based phenotype definitions.

In this study, we present Phecoder, a framework designed to operate directly at the ICD code level. Using pre-trained text embedding models, Phecoder implements semantic search to retrieve and prioritise ICD codes for arbitrary phenotype text queries, allowing users to tailor and expand ICD-based phenotype definitions beyond any fixed catalogue of Phecodes. A central component is an unsupervised ensemble of multiple encoders, based on rank-fusion methods, designed to improve the robustness and completeness of retrieved code lists. We release Phecoder as an open-source, user-friendly Python package that provides both the ensemble retrieval method and a standardised benchmarking framework. This allows researchers to rapidly generate curated ICD shortlists for any phenotype of interest and to efficiently evaluate emerging text embedding models, ensuring that Phecoder can evolve in step with advances in natural language processing. Scalable, text-driven ICD code retrieval tools such as Phecoder will help support more complete, transparent, and reproducible phenotyping for EHR-based research.

## Methods

### Study overview

We developed Phecoder, a semi-automated phenotyping framework that uses semantic retrieval with pre-trained text embedding models to map ICD codes to clinically meaningful phenotypes. Given a free text phenotype description (for example, “eating disorders”), Phecoder encodes both the query and ICD code text descriptions into a shared embedding space, computes pairwise cosine similarity between the phenotype embedding and each ICD embedding, and returns a ranked shortlist of top-k ICD codes for expert review.

Figure 1 illustrates this workflow: starting from a textual phenotype query, Phecoder retrieves semantically related ICD codes, optionally aggregates similarity scores across models, and produces a ranked list that clinicians can audit or extend. Because the method operates on code descriptions rather than fixed hierarchies, it is broadly applicable across different EHR systems.

**Figure 1:**
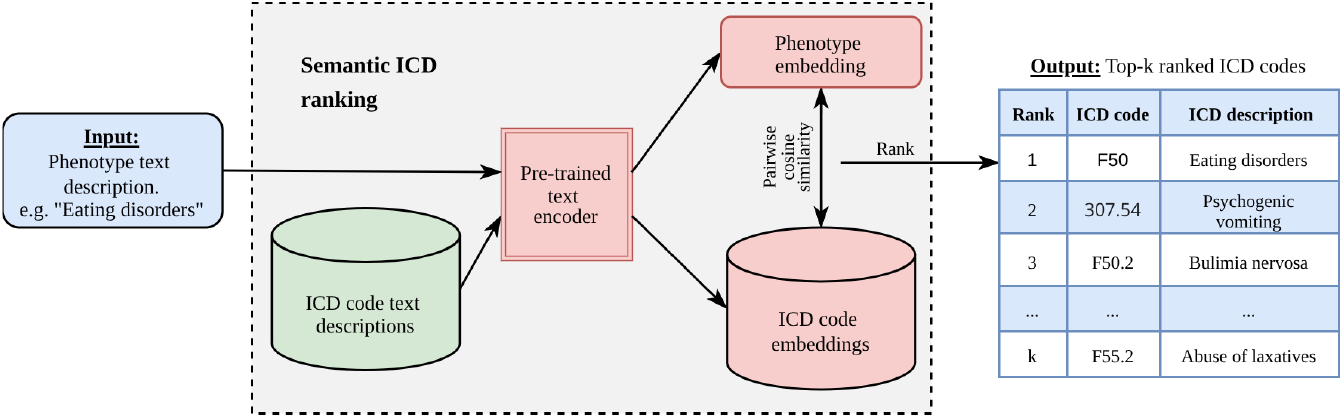
Phecoder semantic retrieval workflow for ICD-based phenotyping. A free-text phenotype description (for example, “Eating disorders”) and all available ICD code text descriptions are encoded into a shared embedding space using a pre-trained text embedding model. Pairwise cosine similarity between the phenotype embedding and each ICD code embedding is then computed to rank codes by semantic relatedness. The system returns a shortlist of the top-k ranked ICD codes for expert review; once approved, these codes can be used to define cohorts for downstream genetic analyses.

The study was structured around three main objectives. First, we evaluated multiple encoders and unsupervised ensemble methods against PhecodeX^8^ as ground truth definitions to assess whether Phecoder can reproduce established ICD-9-CM and ICD-10-CM mappings defined in the reference phenotypes. Second, recognizing that PhecodeX is not an exhaustive ground truth, we used expert clinical review of Phecoder-generated shortlists for six neuropsychiatric case studies to identify additional relevant ICD codes missing from the PhecodeX definition. Third, we examined whether inclusion of these additional ICD codes increased cohort sizes in the Million Veteran Program^21^ (MVP) relative to the original PhecodeX definitions. All analyses undertaken in this study utilized the ICD codes and text descriptions within the MVP.

## Models

We accessed text embedding models via the SentenceTransformers Python package^22^, which provides a unified interface to pretrained transformer-based encoders hosted on the Hugging Face platform^23^. Models were selected to represent top-performing general-purpose encoders^24,25^, EHR-focused models^26^, and baseline SentenceTransformers^27^. Altogether, nine models were selected. More information about the chosen models can be found in Supplementary Table 1. Each model was used to generate a catalog of dense vector embeddings of ICD and PhecodeX text descriptions. Pairwise cosine similarity between embeddings enabled ranking of ICD codes by semantic relatedness to each phenotype definition (Fig. 1).

## Performance evaluation

Performance was evaluated against the PhecodeX reference standard^8^. Phecoder outputs a ranked short list of candidate ICD codes, so evaluation was restricted to the first k positions. For any given Phecode, almost all ICD codes are truly non-relevant. In this imbalanced setting, a trivial model that labels every ICD code as non-relevant would achieve high accuracy while failing to retrieve any relevant codes. Performance was therefore evaluated using recall and average precision at top-k (R@k and AP@k, respectively; see, for example, Manning et al^28^). R@k quantifies the completeness of the Phecode definition by measuring the proportion of relevant ICD codes recovered within this shortlist. We use R@k as the primary performance measure because missing relevant codes would lead to under-ascertainment and smaller, biased cohorts. AP@k complements recall by summarising both the presence and the ordering of relevant ICD codes within the top-k; it is rank-aware and rewards early retrieval of relevant codes.

PhecodeX definitions varied widely in complexity as reflected by the number of included ICD codes (Supplementary Figure 1A). The majority of Phecodes (approximately 66%) contained 10 or fewer ICD codes, rendering their retrieval trivial (median R@10 = 1.0; Supplementary Figure 1B) due to the strong lexical overlap between their ICD descriptions and the Phecode query. To ensure a meaningful evaluation of semantic retrieval, we focused on non-trivial Phecodes, defined as those comprising more than 10 ICD codes. Among the remaining 1,125 Phecodes, ICD code counts were right skewed (median 28, IQR 18–59), meaning that although most phenotypes contained a modest number of ICD codes, a minority encompassed substantially larger sets. Because of this skewed distribution, retrieval performance was summarised using the median across Phecodes rather than the more commonly used mean^28^. The same distribution informed the choice of retrieval depth: selecting k = 100 provided sufficient capacity for the full set of ICD codes to be retrieved for nearly all Phecodes (i.e., R@100 could reach 1.0), while aligning with the practical constraints of expert review, for which a 100 code shortlist is manageable. Note that not all ICD codes listed in PhecodeX existed within the MVP; hence, the ground truth used for a given Phecode was adjusted to be the intersection between the PhecodeX list and the available catalog of ICD codes from the MVP. This ensured all ICD codes were retrievable.

Because different embedding models may encode distinct semantic biases and domain emphases, we first assessed whether their top-ranked ICD retrievals were overlapping or complementary. We quantified pairwise model similarity using the Jaccard Index within the top-100 ranks (J@100), computed separately for (i) ground-truth ICD codes (as defined by PhecodeX) and (ii) candidate ICD codes retrieved for each Phecode. To leverage complementary retrieval signals where present, we constructed several unsupervised ensembles using rank-fusion methods (Supplementary Material).

These approaches require no supervision or hyperparameter tuning and preserve the interpretability and general applicability of the framework.

### Expert clinical review

PhecodeX was used as the primary reference standard for evaluating Phecoder, as it provides the most comprehensive and rigorously curated mapping available. However, PhecodeX does not constitute an exhaustive ground truth^8^. To assess whether Phecoder retrieved clinically plausible ICD codes beyond existing curated definitions, we conducted a proof-of-concept clinical audit using expert review.

We selected six neuropsychiatric Phecodes as case studies: MB_280.11 (Alcohol abuse and dependence), MB_280.2 (Opioid use disorders), MB_286.1 (Bipolar disorder), MB_287.1 (Schizophrenia), MB_293 (Eating disorders), and MB_296 (Personality disorders). For each Phecode, Phecoder was run using all individual models as well as the best-performing ensemble. Retrieved ICD codes were truncated using a PPV threshold of 50% relative to the PhecodeX reference set, after which all ICD codes present in PhecodeX were added to ensure that no known mappings were omitted during review. The combined and de-duplicated list of ICD codes was then randomly shuffled and provided to a clinician with specialist expertise in neuropsychiatric disorders. The expert clinician annotated each ICD code using a three-level ordinal relevance scale (0 = not relevant; 1 = possibly relevant, indicating clinical proximity to the phenotype; 2 = relevant, indicating clear correspondence to the target phenotype). We defined a “strict” phenotype definition as the set of ICD codes assigned 2, and the “extended” phenotype definition as the set of ICD codes assigned 1 or 2. Formal multi-reviewer adjudication and inter-rater reliability assessment were not the objective of this proof-of-concept clinical audit and represent important directions for future work.

### Diagnostic counts in the MVP

For each Phecode case study, we constructed several ICD sets to define patient cohorts within the MVP, including the PhecodeX reference set, and reviewer-defined strict and extended sets retrieved by Phecoder. For every ICD set, we extracted all corresponding diagnosis records for MVP participants. Diagnosis timestamps were collapsed to the calendar day so that multiple codes from the same ICD set recorded on the same day were treated as a single event. For each individual and ICD set, we then counted the number of distinct days on which any ICD code from that set was recorded. These day-level counts were linked to sex and genetic ancestry (HARE^29^) variables, and the distribution of counts was summarised within each subgroup for every ICD set. Note, the “sex” variable was used from a database field previously labeled “gender” in earlier MVP data releases.

## Results

### Single-model performance on PhecodeX ground truth

To establish a performance baseline, we first evaluated how well individual embedding models could reproduce manually curated PhecodeX mappings. This benchmark provides a reference for assessing whether semantic retrieval alone can recover established ICD-9-CM and ICD-10-CM definitions. For each Phecode, 67,632 ICD-9/10 codes from the MVP were ranked according to the semantic similarity of their text descriptions, and performance was assessed against ground-truth mappings. Overall, Qwen-Embedding-4B achieved the highest median recall (Fig. 2A; median R@100 = 0.86), outperforming both clinically trained models such as BioLORD (median R@100 = 0.81) and the larger general purpose model, Qwen-Embedding-8B (median R@100 = 0.85), despite having four billion fewer parameters (though Qwen-Embedding-4B showed slightly lower early-rank precision than Qwen-Embedding-8B; median AP = 0.54 vs 0.56). Performance varied by clinical domain (Fig. 2B), with strongest retrieval observed for “Pregnancy”, “Gastrointestinal”, and “Blood/Immune” Phecodes (median R@100 = 0.97, 0.91, and 0.91, respectively), and weakest performance for “Symptoms”, “Genitourinary”, and “Musculoskeletal” categories (median R@100 = 0.74, 0.76, and 0.76, respectively).

**Figure 2:**
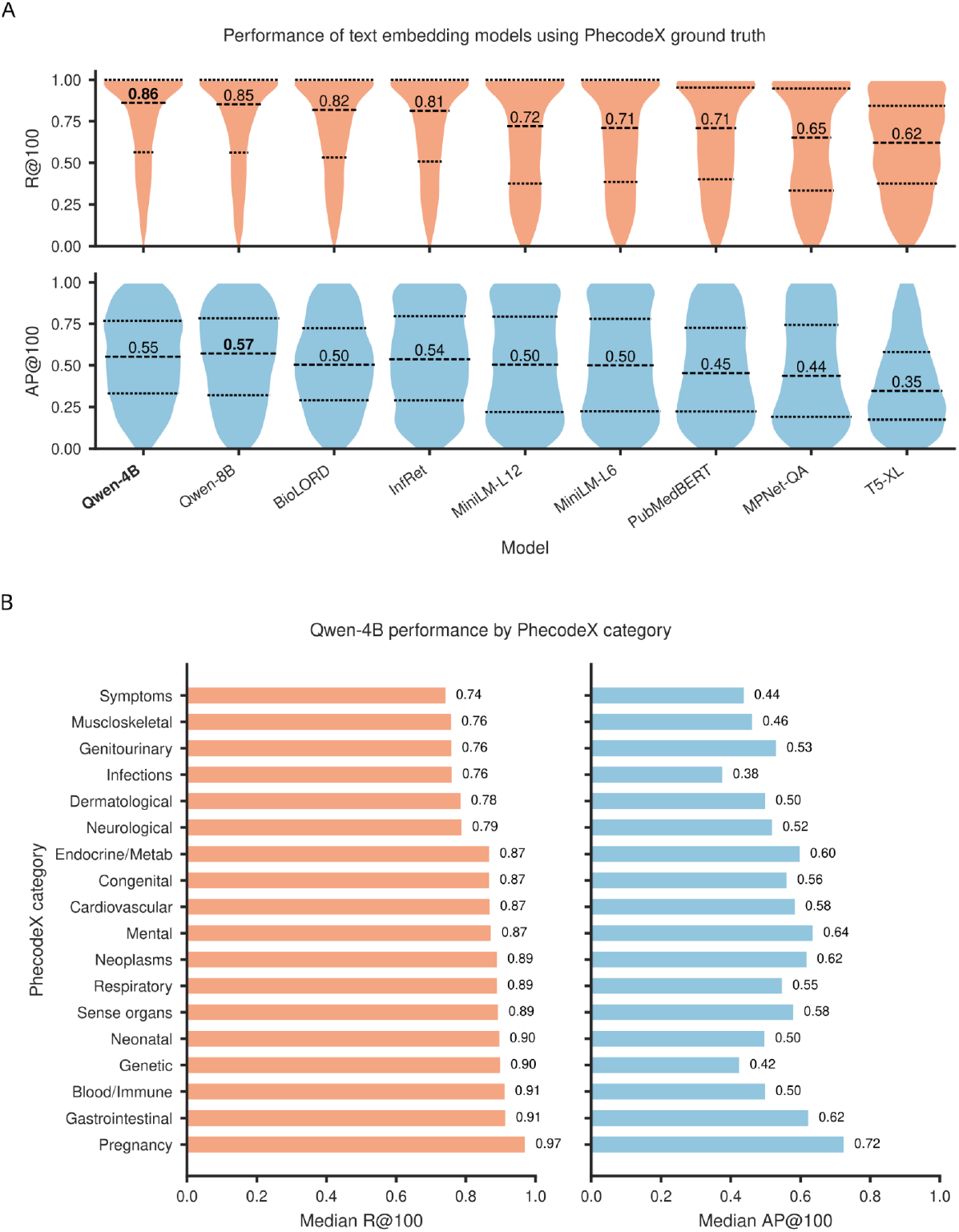
Performance of text embedding models against PhecodeX ground truth. (A) Violin plots show the distribution of the primary recall at top-100 metric (R@100, top), which was used as the main criterion to judge model performance, and the complementary average precision at top-100 (AP@100, bottom) across 1,125 PhecodeX phenotypes for nine text embedding models. Dashed lines mark the median and upper/lower quartiles. Qwen-Embedding-4B (Qwen-4B) achieved the highest median R@100 while maintaining AP@100 comparable to the larger Qwen-Embedding-8B model, outperforming the remaining models. (B) Bar plots show Qwen-Embedding-4B performance stratified by PhecodeX category, with median R@100 (left) and median AP@100 (right) for each domain.

### Model diversity and overlap in retrieval behavior

Because embedding models differ in architecture, training corpus, and clinical specialization, they may capture distinct aspects of ICD semantics. Quantifying their overlap helps determine whether models provide redundant or complementary information. Pairwise model comparisons revealed diversity in retrieval behaviour (Fig. 3). Models showed substantially greater overlap when recovering PhecodeX ground truth ICD codes (median J@100 = 0.64–0.94) than when retrieving candidate ICD codes (median J@100 = 0.15–0.50), indicating that different model architectures prioritize distinct regions of the ICD semantic space. The strongest concordance occurred within model families, consistent with shared or closely related training corpora, but even these closely related models diverged markedly for candidate ICDs. MiniLM-L6 and MiniLM-L12 showed near identical recovery of ground truth ICDs (median J@100 = 0.94) yet only moderate agreement for candidate codes (median J@100 =0.50), while the two best performing models, Qwen-Embedding-4B and Qwen-Embedding-8B, also overlapped strongly for ground truth ICDs (median J@100 =0.88) but agreed only modestly for candidate ICDs (median J@100 =0.40). The least overlap was observed for BioLORD and T5-XL (median J@100 = 0.65 and 0.15 for ground truth and candidate ICDs, respectively), reflecting limited alignment with the broader set of models. Taken together, these findings suggest that individual models capture only partial and often complementary semantic signals, motivating the use of ensemble rank fusion approaches to integrate their strengths and improve robustness when expanding phenotype definitions.

**Figure 3:**
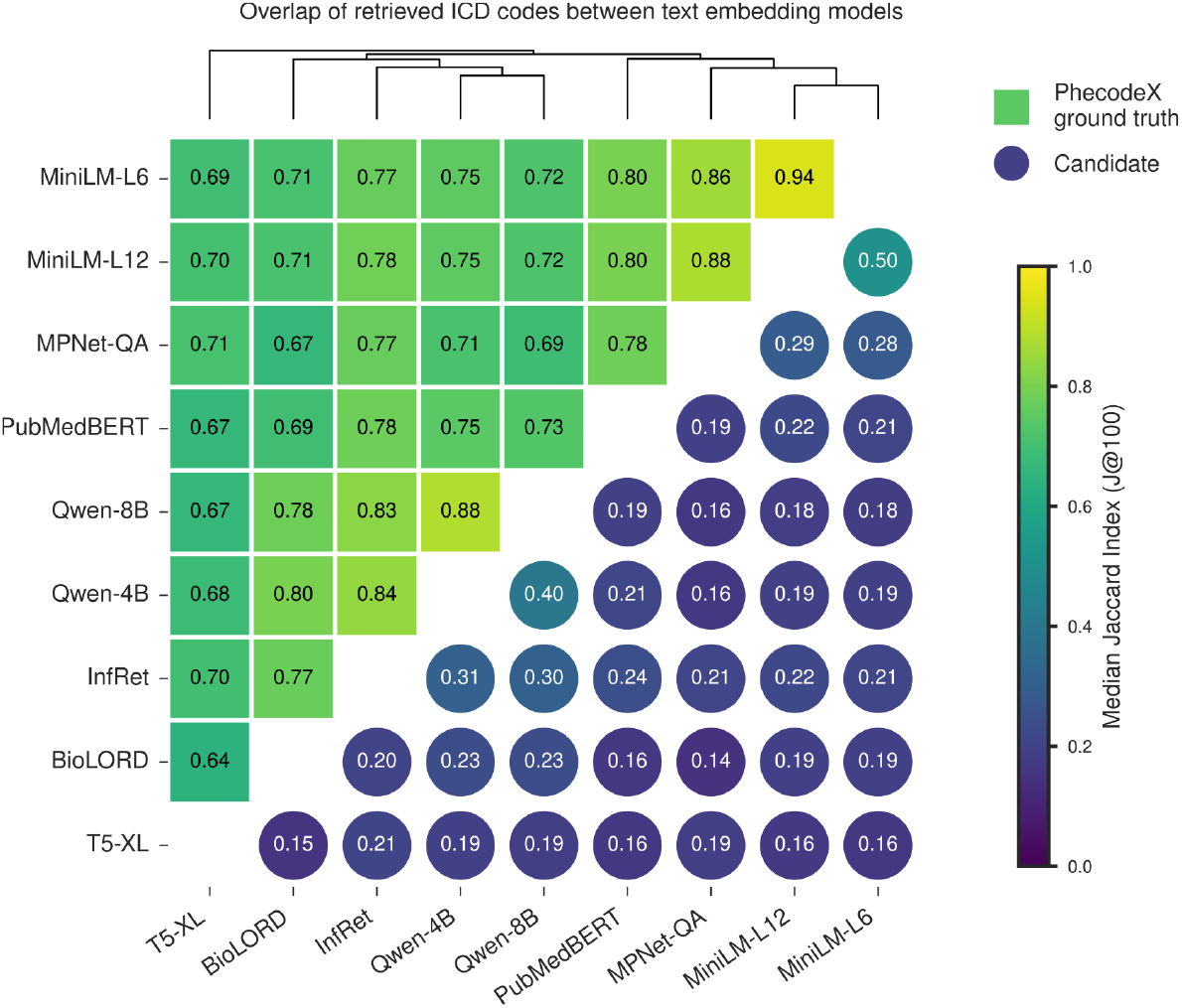
Overlap of retrieved ICD codes between text embedding models. Heatmap showing pairwise similarity between models, quantified as the median Jaccard index of ICD code sets within the top 100 ranks (J@100) across Phecodes. Squares in the upper-left represent overlap of PhecodeX ground-truth ICD codes, and circles in the lower-right represent overlap of candidate ICD codes retrieved beyond PhecodeX; colors indicate the magnitude of J@100. Models are ordered by hierarchical clustering using average-linkage on a distance matrix defined as 1 minus the mean of ground-truth and candidate J@100 values, and the corresponding dendrogram is displayed above the heatmap.

### Ensemble rank-fusion improves retrieval completeness

Given that individual models emphasized different semantic regions, we next tested whether combining their similarity scores could improve retrieval completeness and robustness. Overall, score-level rank fusion methods (Z-Sum, CombSum, Fisher, CombMNZ) achieved greater performance gains than the remaining rank-based approaches (Fig. 4A), which sometimes decreased recall (Mean rank decreased median R@100 by 0.06 vs the best single model). Of the score level methods considered, the Z-sum method, which sums Z-score normalised similarity scores across models, improved recall by three percentage points relative to the best individual model (median R@100 = 0.89 vs 0.86; Fig. 4A) and improved average precision by eight percentage points (median AP@100 = 0.63 vs 0.55), indicating superior early retrieval. CombSum was close behind with equivalent recall (median R@100 = 0.89), but slightly decreased average precision (median AP@100 = 0.62).

**Figure 4:**
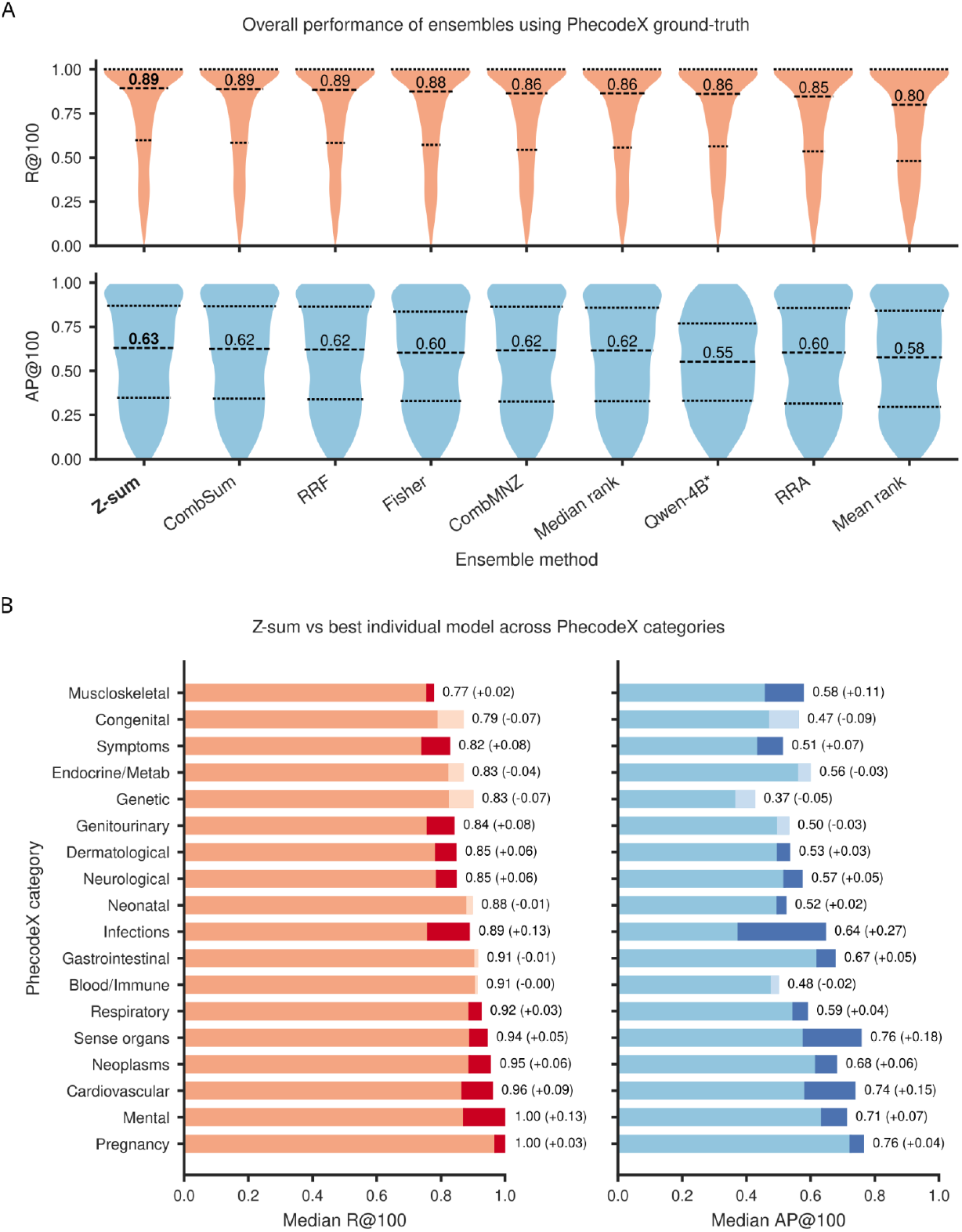
Performance evaluation of ensembles using PhecodeX as the ground truth. (A) Violin plots summarize recall at top-100 (R@100, top) and average precision at top-100 (AP@100, bottom) across 1,125 PhecodeX phenotypes for each ensemble method and for the best single model (starred). Dashed lines mark the median and upper/lower quartiles. Among all ensemble approaches, Z-sum was the best-performing method, achieving the highest median R@100 and therefore the most complete retrieval of ground-truth ICD codes. It also showed the strongest gains in AP@100. (B) For each PhecodeX category, bar plots show median R@100 (left) and median AP@100 (right) for the Z-sum ensemble, with dark/light segments indicating positive/negative change relative to the best individual model. Z-sum exceeds the performance of the best single model across most clinical domains, with particularly pronounced gains in AP@100, indicating a higher concentration of relevant ICD codes at the top of the ranking.

At the category level, Z-sum outperformed the best single model across 12 out of the 18 domains (Fig. 4B). Notably, Z-sum achieved perfect median recall for the “Mental” category (median R@100 = 1.0 vs 0.87; median AP@100 = 0.71 vs 0.64), indicating that the ensemble consistently recovered all relevant ICD codes for the majority of neuropsychiatric phenotypes. This level of completeness was not achieved by any individual model and underscores the value of ensemble methods for semantically diffuse clinical domains. The largest performance increase was observed for the “Infections” category (median R@100 = 0.89 vs 0.76; median AP@100 = 0.64 vs 0.38). The largest performance decrease was observed for the “Genetic” category (median R@100 = 0.83 vs 0.90; median AP@100 = 0.37 vs 0.42), but in absolute terms, this performance was still strong. Thus, Phecoder’s Z-sum ensemble enabled the retrieval of established PhecodeX mappings; the next step was to evaluate its ability in identifying additional ICD codes beyond the existing ground truth.

### Expert review confirms additional ICD codes beyond PhecodeX

While high recall against PhecodeX establishes reproducibility, a central question is whether Phecoder can recover valid codes missing from existing definitions. To test this, we conducted a blinded expert review of six neuropsychiatric case studies. Clinical evaluation demonstrated that Phecoder retrieved clinically meaningful ICD codes absent from PhecodeX. Across all phenotypes, positive predictive value (PPV) curves derived from the strict reviewer labels were consistently greater than or equal to those based on PhecodeX (Fig. 5; first row; orange curve), except for a few ranks in “Personality disorders” where PPV was only marginally lower for a small fraction of ranks. Under the extended reviewer labels (of which the strict reviewer labels are a subset), PPV remained identically 1 from the early to mid ranks for all phenotypes and declined only slightly at later ranks (Fig. 5; first row; green curve): across all phenotypes, the minimum PPV observed across all ranks was 0.88 (Bipolar disorder). These PPV profiles indicated that Phecoder retrieved additional ICD codes (from those defined in PhecodeX phenotypes) that were either directly clinically relevant, or more broadly related, to the phenotype (Fig. 5; second row; orange and green, respectively). For example, in “Opioid use disorders” there were 17 additional ICD codes identified under the strict reviewer definition (Fig. 5; second row; orange curve) representing approximately 15% of the total relevant ICD codes retrieved (Fig. 5; third row; orange curve). All of these codes were “out-of-hierarchy” ICD codes (Fig. 5; fourth/fifth rows; orange curve), i.e. none were easily identifiable via manual inspection of hierarchical parent Phecode mappings. They also included ICD codes with no lexical overlap between text descriptions, e.g. “Heroin causing adverse effects in therapeutic use” (E935.0, ICD-9-CM), demonstrating the advantages of semantic matching. Phecoder retrieved additional ICD codes for all phenotypes under both strict and extended definitions, and full lists can be found in the Supplementary Data.

**Figure 5:**
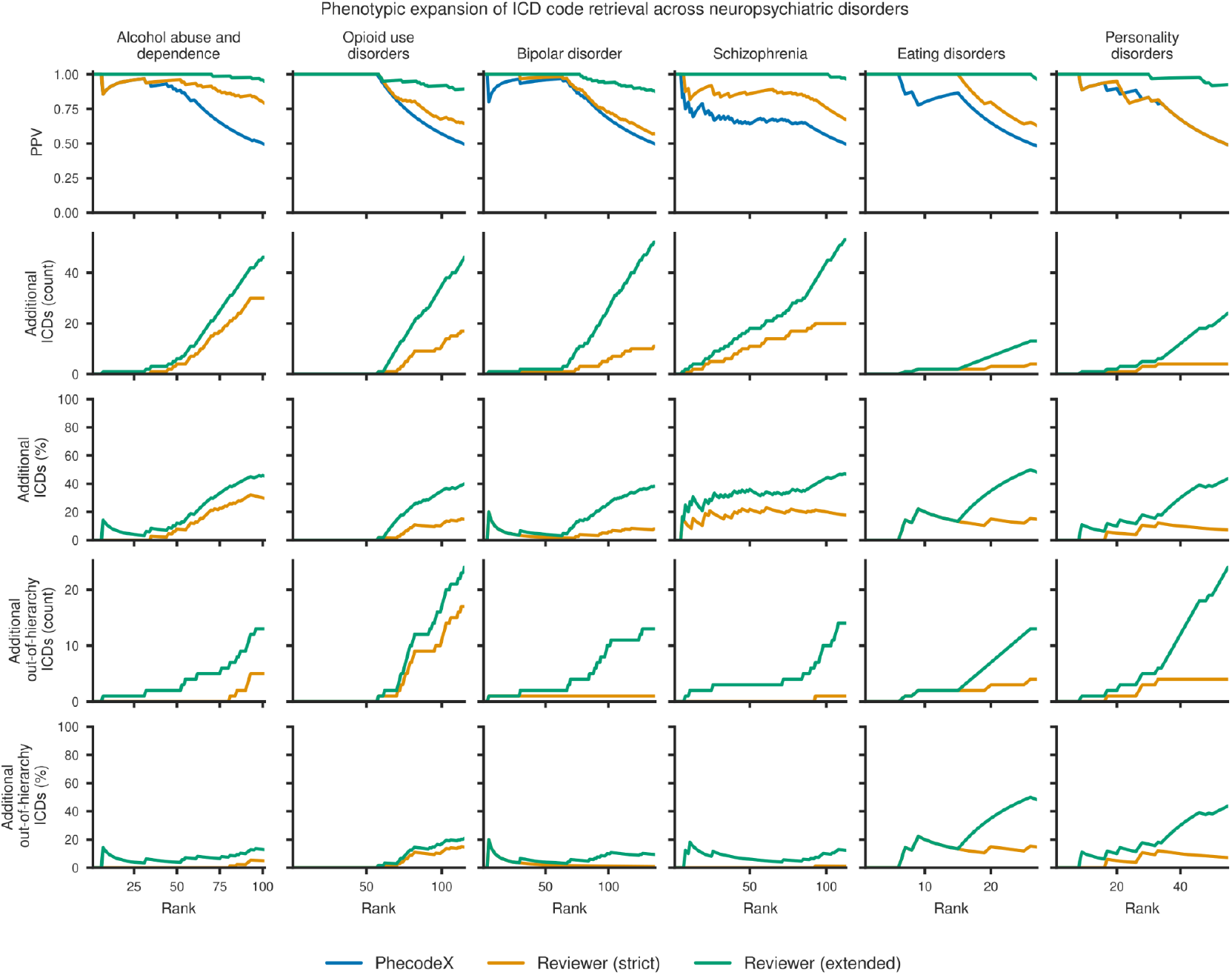
ICD code expansion for six neuropsychiatric phenotypes. Line plots show Phecoder performance, as determined by expert review of ICD shortlists defining “strict” (orange) and “extended” (green) labels, against PhecodeX labels (blue) for six neuropsychiatric case study Phecodes. Rows show: (1) positive predictive value (PPV) across retrieval ranks (cut-off rank was calculated at 50% PPV according to PhecodeX); (2–3) cumulative counts and percentage of additional ICD codes identified; and (4–5) cumulative counts and percentages of additional out-of-hierarchy ICD codes (i.e., those not readily identifiable from parent Phecode mappings). Strict and extended labels reveal that Phecoder retrieves clinically relevant ICD codes beyond those in PhecodeX, including substantial numbers of out-of-hierarchy codes for several phenotypes.

**Figure 6:**
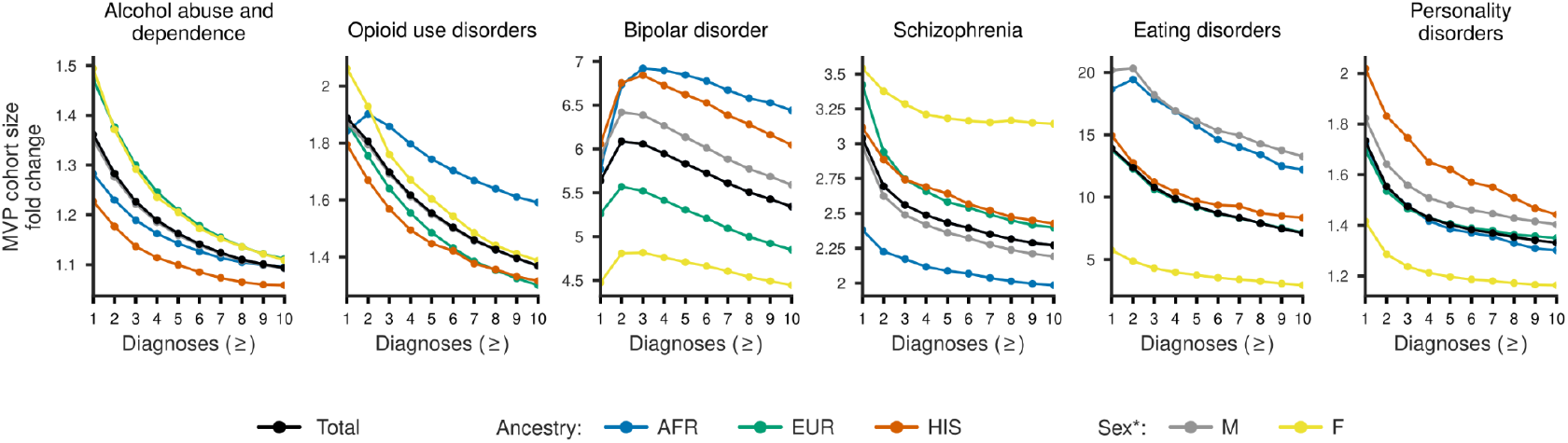
Potential cohort expansion under extended ICD definitions for six neuropsychiatric phenotypes in the Million Veteran Program (MVP). Fold change in MVP cohort size is shown for expert reviewer-defined extended ICD code sets generated by Phecoder, expressed relative to the corresponding PhecodeX definitions, across increasing diagnosis count thresholds. Results are stratified by genetic ancestry (AFR, EUR, HIS) and sex, with overall totals shown in black. The extended definitions yield substantial potential cohort expansion, particularly at lower diagnostic thresholds, with the largest fold increases observed for eating disorders and bipolar disorder across several demographic groups. *Note, the “sex” variable was used from a database field previously labeled “gender” in earlier MVP data releases.

### Cohort expansion in the MVP

Finally, to determine whether these expanded definitions translate into practical analytic gains, we examined their impact on cohort ascertainment within MVP. Using the expert-reviewed extended phenotype definitions, Phecoder-based ICD code shortlists produced consistent increases in MVP cohort sizes across all six neuropsychiatric case studies, with median fold changes of approximately two-fold across diagnosis count thresholds (IQR ≈ 1.4 – 5.3; Sup. Fig. 2A). Cohort expansion was greatest at lower diagnostic count thresholds and diminished as stricter inclusion criteria were applied. “Eating disorders” showed the most pronounced increases, particularly in male (13.3 – 20.3 fold) and African ancestry (12.2 – 19.4 fold) cohorts. “Bipolar disorder” demonstrated the greatest increases in African (5.8 – 6.9 fold) and Hispanic ancestries (6.0 – 6.8 fold), whereas “Schizophrenia” showed the greatest increases among female (3.1 – 3.5 fold), European ancestry (2.4 – 3.4 fold) and Hispanic ancestry (2.4 – 3.1 fold) cohorts. More modest but still consistent increases were observed for “Personality disorders” (1.2 – 2.0 fold), “Opioid use disorders” (1.3 – 2.1 fold), and “Alcohol abuse and dependence” (1.1 – 1.5 fold). Under the strict phenotype definitions, cohort expansion was more limited overall, with the median fold change close to one (IQR ≈ 1.0 – 1.4; Sup. Fig. 2B), but remained substantial for schizophrenia (1.7 – 2.4 fold; Sup. Fig. 3) and eating disorders (1.3 – 4.5 fold). Cohort sizes were consistently larger than in the corresponding PhecodeX definitions (fold change > 1) across strict/extended phenotype definitions, demographic strata (sex and ancestry), and diagnosis count thresholds.

## Discussion

Phecoder is an automated framework that uses semantic search with pre-trained text embedding models to retrieve and prioritize ICD codes for arbitrary phenotype text queries. A major advantage of Phecoder is that it does not rely on a fixed phenotype list: any phenotype that can be described in text can be encoded, and ICD codes retrieved. This enables “continuous phenotyping”, whereby ICD-based definitions can be dynamically refined as new evidence, coding systems, or analytic goals emerge. To our knowledge, this is the first systematic benchmarking of text embedding models for ICD-based phenotyping using PhecodeX as a ground-truth. We showed that Phecoder can recover PhecodeX mappings with high sensitivity using an unsupervised rank fusion ensemble method. The ensemble improved recall in 12 of the 18 clinical domains considered (including perfect median sensitivity in the “Mental” health domain) and substantially enhanced early retrieval, increasing the concentration of relevant ICD codes among the highest ranked positions and reducing the number of codes that investigators must manually review. Across six neuropsychiatric case studies in the MVP, Phecoder-expanded phenotypes increased cohort sizes by a median of approximately two-fold, with some phenotypes showing expansions exceeding an order of magnitude. For practitioners, this demonstrates that semantic retrieval can uncover large pools of clinically relevant but previously unmapped patients while keeping the manual review burden manageable.

Phecoder enables both direct cohort expansion through omitted diagnostic codes and indirect expansion by surfacing clinically adjacent codes that can support downstream modeling.Beyond cohort expansion, text-driven tools like Phecoder will become increasingly important as diagnosis code systems evolve. ICD-11^30^ is already in use in many countries and introduces new structures that make manual construction of Phecode-like mappings increasingly challenging. Because Phecoder operates on code descriptions rather than fixed hierarchies, it is naturally extensible to ICD-11 and can assist in building crosswalks between ICD-9, ICD-10, and ICD-11. In principle, the same framework can support future iterations of PhecodeX by proposing candidate ICD-11 mappings and refining those of ICD-9/10.

Several limitations of this study should be acknowledged. First, the cohort sizes reported for strict and extended phenotypes are intended to illustrate the magnitude of untapped patient numbers rather than to estimate true case counts; in practice, final cohort sizes will be smaller once study specific inclusion criteria are applied. Second, our evaluation of cohort expansion was restricted to the MVP and a limited set of neuropsychiatric case studies; broader assessments across other populations, health systems, and disease areas are needed.. Third, the clinical review was designed as a proof-of-concept audit using a single expert reviewer; future work incorporating multi-reviewer adjudication will be important for assessing inter-rater variability. We also focused exclusively on a subset of open-source text embedding models. While more advanced proprietary models exist, the strong performance observed here suggests that open-source approaches provide an effective and transparent foundation for ICD-based phenotyping. Taken together, this work demonstrates how reproducible, text-driven phenotyping can function as foundational infrastructure for EHR-based analyses, supporting scalable and transparent cohort definition while preserving clinical interpretability.

## Code availability

The Phecoder package is available at GitHub: http://www.github.com/DiseaseNeuroGenomics/Phecoder

