## Supplementary material for "Phecoder: semantic retrieval for auditing and expanding ICD-based phenotypes in EHR biobanks": Suplemental Material

**Affiliations:**
[1]Department of Psychiatry, Icahn School of Medicine at Mount Sinai, New York, NY, USA
[2]Center for Disease Neurogenomics, Icahn School of Medicine at Mount Sinai, New York, NY USA
[3]Friedman Brain Institute, Icahn School of Medicine at Mount Sinai, New York, NY, USA
[4]Department of Genetics and Genomic Science, Icahn School of Medicine at Mount Sinai, New York, NY, USA
[5]Mental Illness Research, Education, and Clinical Center (VISN 2 South), James J. Peters VA Medical Center, Bronx, NY, USA
[6]Center for Precision Medicine and Translational Therapeutics, James J. Peters VA Medical Center, Bronx, NY, USA.
[7]Department of Artificial Intelligence and Human Health, Icahn School of Medicine at Mount Sinai, New York, NY, USA

**Ensemble methods** **2**
**Supplementary tables** **3**
**Supplementary figures** **4**
**References** **7**
**VA Million Veteran Program Core Acknowledgements** **8**

#### Ensemble methods

We evaluated several standard unsupervised rank- and score-based ensemble methods to combine retrieval outputs across text embedding models. All methods operate independently for each Phecode and fuse candidate ICD codes based on their ranks or cosine similarity scores. Exact implementations of these methods are available in the Phecoder GitHub repository referenced in the main text.

Rank-based fusion methods combined ICD-code ranks across models to derive a consensus ordering. Reciprocal Rank Fusion (RRF) transformed ranks using a reciprocal function which were summed across models. Mean Rank (sometimes referred to as the Borda count) averaged ranks across models, while Median Rank used the median position to provide robustness to outlier models. Robust Rank Aggregation (RRA) converted observed ranks into p-values reflecting their deviation from random expectation and aggregated these into a Z-score, prioritizing ICD codes repeatedly appearing near the top of model-specific rankings.

Score-based fusion methods combined similarity scores rather than ranks. Z-sum standardized scores within each model and summed the resulting Z-scores, highlighting ICD codes strongly supported across encoders. CombSum applied min–max normalization to scores within each model before summing them, and CombMNZ multiplied this sum by the number of models contributing a valid score, rewarding consistent model agreement. Fisher's method converted similarity scores into empirical p-values and combined them using the Fisher statistic, yielding stronger evidence for ICD codes supported across multiple models.

#### Supplementary tables

| **Model name (Hugging Face)** | **Abbreviated model name** | **Clinical focus?** | **Number of parameters** | **Reason for inclusion** |
|---|---|---|---|---|
| FremyCompany/BioLORD-2023[1] | BioLORD | Yes | ~110M | Clinical domain–specialised encoder trained on biomedical and clinical text. |
| infly/inf-retriever-v1[2] | InfRet | No | ~7B | Top performing dense retriever on medical domain retrieval benchmarks. (MTEB)[3]. |
| sentence-transformers/all-MiniLM-L6-v2[4] | MiniLM-L6 | No | ~23M | Efficient general-purpose embedder with strong performance for its size. |
| sentence-transformers/sentence-t5-xxl[4] | T5-XL | No | ~5B | Unique T5-based model[5] offering high quality semantic similarity representations. |
| sentence-transformers/multi-qa-mpnet-base-dot-v1[4] | MPNet-QA | No | ~110M | Top performer from original sentence-transformer[4] models for semantic retrieval. |
| sentence-transformers/all-MiniLM-L12-v2[4] | MiniLM-L12 | No | ~33M | Efficient general-purpose embedder with strong performance for its size. Slightly larger model than the other MiniLM model considered. |
| NeuML/pubmedbert-base-embeddings[6] | PubMedBERT | Yes | ~110M | Clinical domain–specialised. Trained PubMed[7] title–abstract pairs. |
| Qwen/Qwen3-Embedding-8B | Qwen-8B | No | ~8B | Included as a state-of-the-art large embedding model with leading performance on semantic similarity and retrieval benchmarks[3]. |
| Qwen/Qwen3-Embedding-4B | Qwen-4B | No | ~4B | Selected as a smaller, computationally lighter variant of the 8B parameter model |

**Supplementary Table 1:** Summary of individual text embedding models considered in this study. More details can be found in the referenced papers, or searching for the relevant Hugging Face repositories (the Hugging Face repository name is the same as the Model name).

#### Supplementary figures

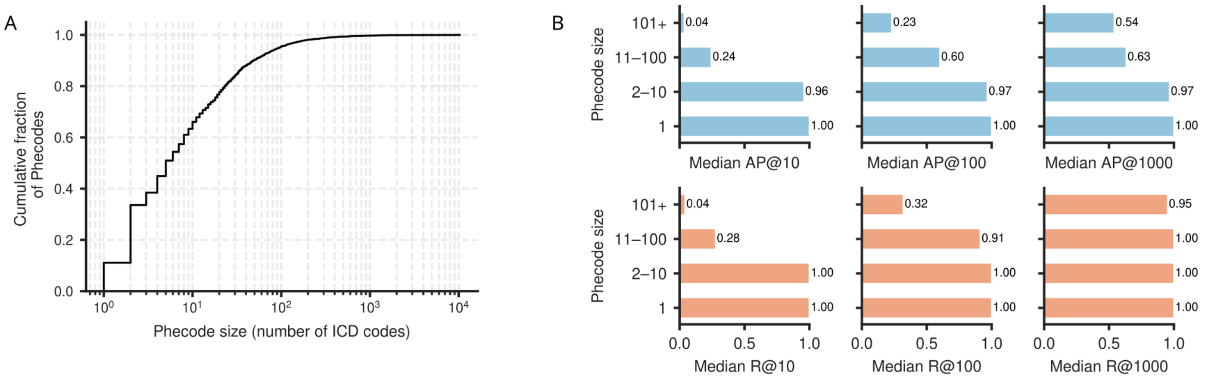


**Supplementary Figure 1: Distribution of Phecode sizes and retrieval performance as a function of phenotype complexity.** (A) Cumulative distribution of PhecodeX phenotype sizes, expressed as the number of ICD codes per Phecode. Most Phecodes contain ten or fewer ICD codes, with a long right tail reflecting phenotypes of substantially greater complexity. (B) Median average precision (median AP@k; top row) and median recall (median R@k; bottom row) at k = 10, 100, and 1,000, stratified by Phecode size. Retrieval performance is near-perfect for small Phecodes while more complex phenotypes show lower performance, motivating the focus on non-trivial Phecodes (>10 ICD codes) in the main evaluation.

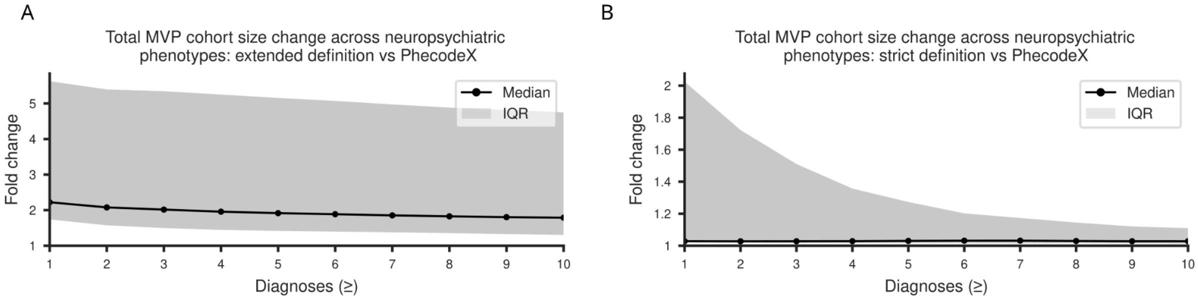


**Supplementary Figure 2: Overall potential cohort expansion across six neuropsychiatric phenotypes in the Million Veteran Program.** (A) Fold change in total MVP cohort size using extended reviewer-defined ICD code sets generated by Phecoder, expressed relative to PhecodeX, across increasing diagnosis-count thresholds. The curve shows the median fold change across the six neuropsychiatric phenotypes, with the shaded region indicating the interquartile range (IQR). (B) Corresponding fold-change for the strict reviewer-defined ICD code sets. Cohort expansion is consistently larger under the extended definitions, with both definitions showing the greatest relative increase at lower diagnostic thresholds.

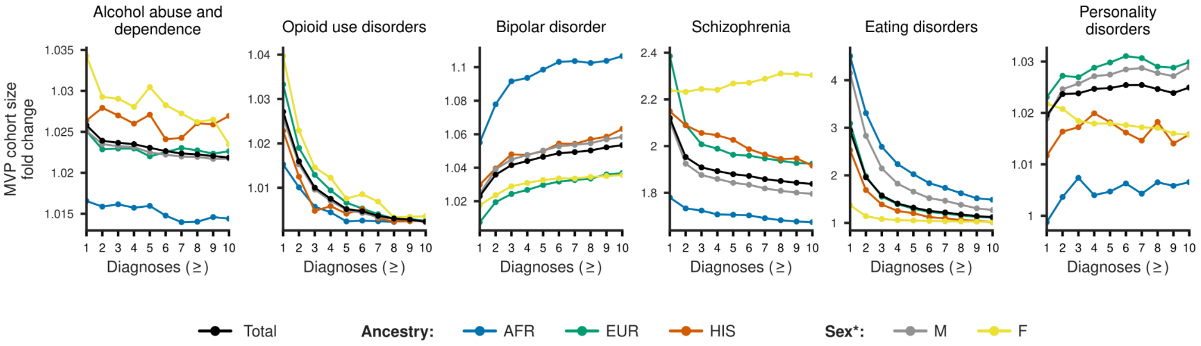


**Supplementary Figure 3:** Potential cohort expansion under strict ICD definitions for six neuropsychiatric phenotypes in the Million Veteran Program. Fold change in MVP cohort size is shown for expert reviewer-defined strict ICD code sets generated by Phecoder, expressed relative to the corresponding PhecodeX definitions, across increasing diagnosis count thresholds. Results are stratified by genetic ancestry (AFR, EUR, HIS) and sex, with overall totals shown in black. Cohort expansion under the strict definitions is more limited than under the extended definitions but remains evident for several phenotypes, notably schizophrenia and eating disorders, particularly at lower diagnostic thresholds. *Note, the “sex” variable was used from a database field previously labeled “gender” in earlier MVP data releases.

### VA Million Veteran Program Core Acknowledgements


**MVP Program Office**
- Sumitra Muralidhar, Ph.D., Program Director
US Department of Veterans Affairs, 810 Vermont Avenue NW, Washington, DC 20420
- Jennifer Moser, Ph.D., Associate Director, Scientific Programs
US Department of Veterans Affairs, 810 Vermont Avenue NW, Washington, DC 20420
- Jennifer E. Deen, B.S., Associate Director, Cohort & Public Relations
US Department of Veterans Affairs, 810 Vermont Avenue NW, Washington, DC 20420

**MVP Steering Committee**
- Co-Chair: Philip S. Tsao, Ph.D.
VA Palo Alto Health Care System, 3801 Miranda Avenue, Palo Alto, CA 94304
- Co-Chair: Sumitra Muralidhar, Ph.D.
US Department of Veterans Affairs, 810 Vermont Avenue NW, Washington, DC 20420
- J. Michael Gaziano, M.D., M.P.H.
VA Boston Healthcare System, 150 S. Huntington Avenue, Boston, MA 02130
- Adriana Hung, M.D., M.P.H.,
VA Tennessee Valley Healthcare System, 1310 24th Avenue, South Nashville, TN 37212
- Dave Oslin, M.D.
Philadelphia VA Medical Center, 3900 Woodland Avenue, Philadelphia, PA 19104
- Deepak Voora, M.D.
Durham VA Medical Center, 508 Fulton Street, Durham, NC 27705

**MVP Co-Principal Investigators**
- J. Michael Gaziano, M.D., M.P.H.
VA Boston Healthcare System, 150 S. Huntington Avenue, Boston, MA 02130
- Philip S. Tsao, Ph.D.
VA Palo Alto Health Care System, 3801 Miranda Avenue, Palo Alto, CA 94304

**MVP Core Operations**
- Jessica V. Brewer, M.P.H., Director, MVP Cohort Operations
VA Boston Healthcare System, 150 S. Huntington Avenue, Boston, MA 02130
- Mary T. Brophy M.D., M.P.H., Director, VA Central Biorepository
VA Boston Healthcare System, 150 S. Huntington Avenue, Boston, MA 02130
- Kelly Cho, M.P.H, Ph.D., Director, MVP Phenomics
VA Boston Healthcare System, 150 S. Huntington Avenue, Boston, MA 02130
- Lori Churby, B.S., Director, MVP Regulatory Affairs
VA Palo Alto Health Care System, 3801 Miranda Avenue, Palo Alto, CA 94304
- Jacob T. Kean, Ph.D., Acting Director, VA Informatics and Computing Infrastructure (VINCI)
MVP Core Acknowledgements for Publications_October 2025
VA Salt Lake City Health Care System, 500 Foothill Drive, Salt Lake City, UT 84148
- Saiju Pyarajan Ph.D., Director, Data and Computational Sciences
VA Boston Healthcare System, 150 S. Huntington Avenue, Boston, MA 02130
- Robert Ringer, Pharm.D., Director, VA Albuquerque Central Biorepository
New Mexico VA Health Care System, 1501 San Pedro Drive SE, Albuquerque, NM 87108
- Luis E. Selva, Ph.D., Director, MVP Biorepository Coordination
VA Boston Healthcare System, 150 S. Huntington Avenue, Boston, MA 02130
- Shahpoor (Alex) Shayan, M.S., Director, MVP PRE Informatics
VA Boston Healthcare System, 150 S. Huntington Avenue, Boston, MA 02130
- Brady Stephens, M.S., Principal Investigator, MVP Information Center
Canandaigua VA Medical Center, 400 Fort Hill Avenue, Canandaigua, NY 14424
- Stacey B. Whitbourne, Ph.D., Director, MVP Cohort Development and Management
VA Boston Healthcare System, 150 S. Huntington Avenue, Boston, MA 02130
